# Altered Unimodal-to-Transmodal Cortical Hierarchy Before Transition to Psychosis in Clinical High-Risk Individuals

**DOI:** 10.64898/2026.08.30.26361747

**Authors:** Yichuan Wang, Enhui Zhang, Shaolei Guo, Anfu Deng, Bihua Xu, Jiaxi Liao, Yulin Wang, Debo Dong

**Author notes:** These authors contributed equally to this work (Yichuan Wang, Enhui Zhang, Shaolei Guo). To whom correspondence should be addressed. **Dr. Yulin Wang**, Faculty of Psychology, Southwest University, Chongqing 400715, China, **Dr. Debo Dong**, Key Laboratory of Cognition and Personality, Ministry of Education, Faculty of Psychology, Southwest University, Chongqing 400715, China, or.

## Abstract

Psychosis has long been conceptualized as a disorder of disrupted hierarchical integration across distributed brain systems, yet it remains unclear whether alterations in macroscale cortical hierarchy are already present before illness onset and are associated with subsequent transition to psychosis. Using connectome gradient mapping, we characterized baseline cortical hierarchical architecture along the unimodal-to-transmodal axis in 580 participants from the NAPLS-3 cohort, including converters (CHR-C, n = 56), non-converters (CHR-NC, n = 434), and healthy controls (HC, n = 90). Group differences were assessed at regional, network, and global levels. Group comparisons revealed that CHR-C individuals, relative to the other two groups, exhibited bidirectional alterations selectively along the sensorimotor-to-association gradient, with reduced values in the visual network alongside elevated values in the default mode network, indicating greater separation between sensory and transmodal systems along the gradient. At the global level, CHR-C showed increased explained variance, range, and variation of this gradient, collectively indicating hierarchical expansion. Notably, greater explained variance of this gradient was associated with a shorter time to conversion to psychosis, while increased gradient range and variation were associated with higher positive symptom severity across CHR individuals. These findings indicate that expansion of the sensorimotor-to-association connectome hierarchy is already present before psychosis onset in individuals who subsequently convert to psychosis. This altered hierarchical organization may reflect greater decoupling between sensory and transmodal systems and may characterize neurobiological changes associated with progression from a clinical high-risk state to psychotic illness.

## Introduction

Psychosis has long been conceptualized as a disorder involving impaired integration across distributed brain systems rather than dysfunction of isolated regions alone [1–3]. In particular, converging theoretical and empirical work suggests that psychotic symptoms may arise from disrupted interactions between lower-order sensory systems and higher-order transmodal association systems, reflecting abnormalities in the hierarchical organization of cortical function [4–7]. This sensory-to-association hierarchy is thought to support the progressive integration of perceptual information with higher cognitive and self-referential processes [8,9], and its disruption may therefore represent a systems-level feature relevant to psychosis.

In recent years, functional connectome gradient mapping has emerged as a useful framework for characterizing the hierarchical organization of the human cortex [10,11]. The human cortex is organized along continuous functional axes, anchored by unimodal sensory and sensorimotor systems at one end and transmodal association systems at the other, through which information is progressively integrated from concrete sensory input to more abstract representations [12,13]. By reducing high-dimensional resting-state functional connectivity patterns into low-dimensional continuous manifolds, gradient approaches capture this macroscale hierarchical architecture and provide a spatially continuous description of cortical organization rather than one defined by discrete network boundaries [14]. In schizophrenia, studies using this framework have reported abnormalities along the unimodal-to-transmodal axis and altered gradient architecture involving both unimodal and transmodal systems, particularly the visual, attention, and default mode networks [15–17], with such alterations also linked to symptom severity, cognition, and functional outcome [18–21]. Together, these findings suggest that disrupted hierarchical organization of the functional connectome may be relevant to psychotic disorders more broadly [22–24].

An important unresolved question, however, is whether these abnormalities are already present before psychosis onset or instead emerge only after illness onset. Most prior gradient studies have focused on established schizophrenia, first-episode schizophrenia, or broad early psychosis samples [5,15–21,25], limiting direct inference about pre-onset disease processes. Notably, one study comparing early psychosis and schizophrenia found more pronounced abnormalities in chronic stage, suggesting that hierarchical dysfunction may become more evident as illness progresses [5]. Nevertheless, this does not rule out the possibility that selective alterations along the unimodal-to-transmodal axis may already be detectable in individuals prior to conversion.

The clinical high-risk for psychosis state provides an opportunity to address this issue. Because only a subset of CHR individuals subsequently transition to psychosis, outcome-defined comparisons between converters and non-converters can help identify neural alterations associated with later illness onset rather than general risk status alone [26]. In the present study, we examined baseline resting-state functional connectome gradients in the clinical high-risk group of the North American Prodrome Longitudinal Study 3 (NAPLS-3) cohort [27]. We specifically tested whether alterations in unimodal-to-transmodal gradient organization are detectable before psychosis onset by comparing CHR participants who later transitioned to psychosis with those who did not, as well as healthy controls. Gradient alterations were characterized at the regional, network, and global levels, with particular interest in systems spanning unimodal and transmodal domains, including the visual network and default mode network. We further examined associations between gradient measures, symptom severity, and time to conversion. We hypothesized that individuals who later transitioned to psychosis would already show baseline abnormalities in unimodal-to-transmodal gradient organization, consistent with a neural alteration that precedes psychosis onset [28].

### Participants

Data for the present study were drawn from NAPLS-3, a large-scale, longitudinal, multisite observational cohort of individuals at clinical high-risk for psychosis (CHR-P) in the United States [27]. The cohort was recruited across nine research centers and included 560 enhanced-risk CHR-P participants, 150 non-enhanced-risk CHR-P participants, and 96 healthy controls, all aged 12–30 years. CHR-P status was established using version 5.6 of the Structured Interview for Psychosis-Risk Syndromes (SIPS). For the present analyses, only participants in the enhanced-risk CHR-P subgroup were included. This restriction was applied because the enhanced-risk subgroup represents a more clinically enriched and relatively homogeneous high-risk population, characterized by greater symptom burden and a higher likelihood of adverse clinical outcomes. Focusing on this subgroup therefore allowed a more targeted investigation of the neurobiological correlates of psychosis risk while reducing heterogeneity associated with broader CHR-P definitions. Classification into the enhanced-risk subgroup required fulfillment of at least one of the following criteria: (i) a score of 4 or higher on either P1 (unusual thought content) or P2 (suspiciousness) of the Scale of Psychosis-Risk Symptoms (SOPS); (ii) a score of 3 on both P1 and P2; or (iii) significant impairment in symbol coding and verbal memory on the Brief Assessment of Cognition in Schizophrenia (BACS), defined as performance at or below the 10th percentile relative to age-adjusted normative data for adolescents or adults.

CHR-P participants were excluded if they had a current or lifetime Axis I psychotic disorder, including affective psychosis, an IQ below 70, a history of central nervous system disease, or psychosis-risk symptoms considered secondary to another Axis I disorder. Healthy controls were required not to meet criteria for any psychosis-risk syndrome and were additionally excluded if they had a current or past psychotic disorder, a Cluster A personality disorder, a first-degree family history of psychosis-spectrum or related disorders, or current psychotropic medication use. During follow-up, conversion to psychosis was defined according to the SIPS Presence of Psychotic Symptoms (POPS) criteria. Specifically, conversion was considered to have occurred when at least one of the five SOPS positive symptoms reached a psychotic level of intensity (score = 6) at a frequency of at least 1 hour per day on 4 days per week during the past month.

The present study examined whether baseline alterations in cortical hierarchy were associated with later transition to psychosis. Resting-state functional MRI (rs-fMRI) data acquired at the initial recruitment visit were analyzed. Only participants with available and usable baseline rs-fMRI data were included in the analyses. Participants were excluded if rs-fMRI data were unavailable or did not pass quality-control procedures, including exclusion due to excessive head motion during fMRI scanning, defined as a mean Framewise Displacement (FD) greater than 0.2 or the proportion of bad data points (FD > 0.5) [29], exceeding 30%. After these exclusions, the final analytical sample consisted of 90 healthy controls (HC), 56 CHR participants who converted to psychosis during follow-up (CHR-C), and 434 CHR participants who did not convert during follow-up (CHR-NC), see Table 1 for sample characteristics. All NAPLS-3 study procedures were approved by the institutional review boards of the participating institutions, and informed consent was obtained from all participants, with parental consent provided when required. This study was also approved by the Ethics Committee of Southwest University.

**Table 1.**
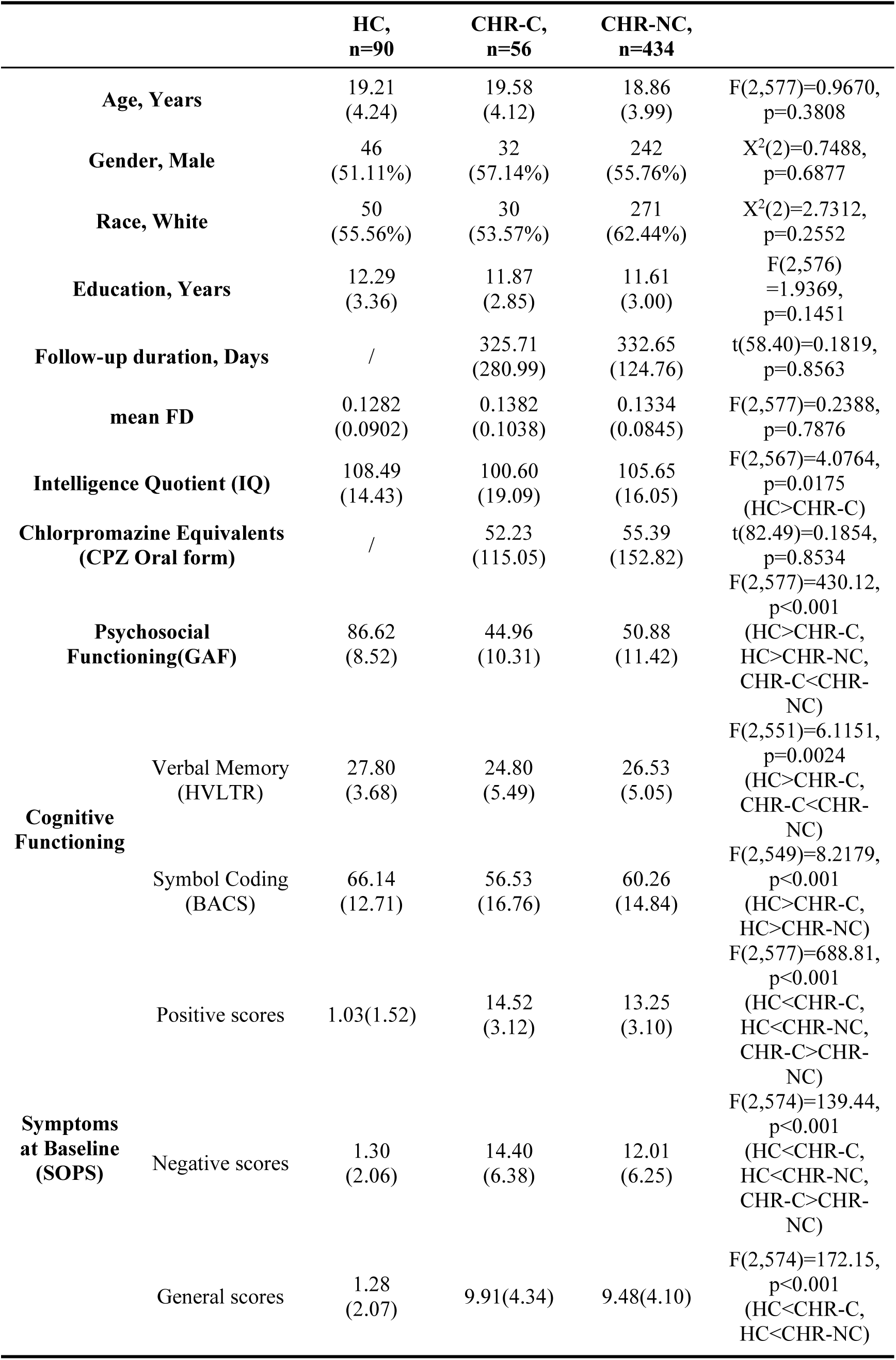
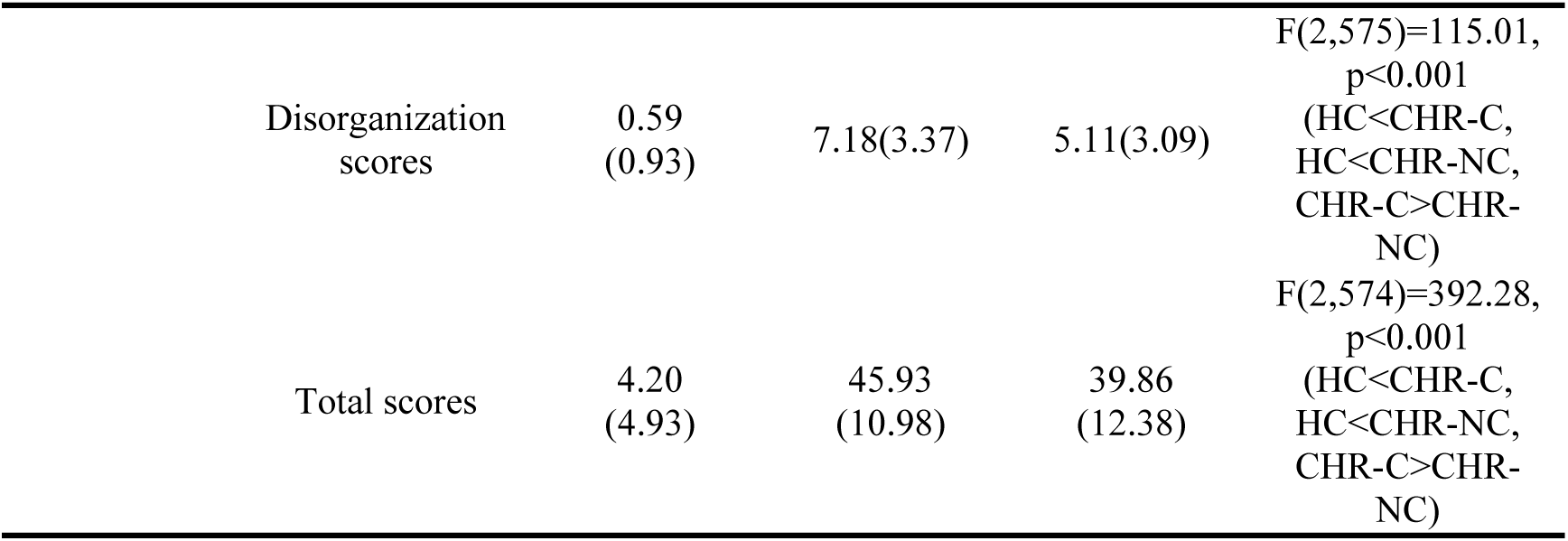
Demographic and Clinical Information.

### MRI Data Acquisition and Processing

Identical MRI acquisition procedures were implemented at all nine participating sites. Cross-site harmonization was facilitated through the use of a structural phantom together with nine traveling human phantoms, each of whom underwent two scans at every site, as detailed previously [27]. In the present study, analyses were based on T1-weighted anatomical images and single-band resting-state functional MRI (rs-fMRI) data.

T1-weighted anatomical images were acquired on 3.0-T Siemens Skyra scanners using a 3D MPRAGE sequence (TR = 2.4 s, TE = 1.96 ms, TI = 1.0 s, flip angle = 8°, slice thickness = 1 mm, matrix = 256 × 256). Resting-state functional MRI data were obtained using a single-band gradient-echo EPI sequence (TR = 2.5 s, TE = 29 ms, flip angle = 85°, slice thickness = 3.5 mm, matrix = 64 × 64).

T1-weighted (T1w) images and resting-state fMRI data were preprocessed using fMRIPrep (version 24.1.1) [30], implemented in Nipype (version 1.8.6). For anatomical preprocessing, T1w images underwent intensity non-uniformity correction, skull stripping, tissue segmentation, and nonlinear spatial normalization to MNI standard space. For functional preprocessing, BOLD reference images were generated for motion correction, and the functional data were co-registered to the corresponding T1w images using boundary-based registration. Head-motion parameters, framewise displacement and other confounding time series were estimated by fMRIPrep. The preprocessed functional images were then spatially smoothed with a 6-mm full-width at half maximum Gaussian kernel, followed by nuisance regression including the Friston 24-parameter motion model and mean signals from white matter and cerebrospinal fluid. Finally, the data were temporally band-pass filtered (0.01–0.1 Hz) to retain low-frequency fluctuations relevant to resting-state functional connectivity.

### Gradient analysis

Regional time series were extracted from 400 cortical parcels defined by the Schaefer atlas [31], which is widely used in cortical functional connectivity analyses and assigns each parcel to one of the seven canonical Yeo functional networks: the visual network, somatomotor network, dorsal attention network, ventral attention network, limbic network, frontoparietal network, and default mode network. Region-wise functional connectivity matrices were then constructed by calculating pairwise Pearson correlation coefficients between the regional time series. The resulting connectivity matrices were Fisher z-transformed prior to further analysis.

To characterize large-scale connectome hierarchical organization, connectome gradients were estimated using diffusion map embedding as implemented in the MATLAB-based BrainSpace toolbox [32]. For each node, only the strongest 10% of functional connections were retained[15,33]. Cosine similarity was then computed between pairs of nodes, and the resulting similarity matrix was converted into a normalized angle matrix to avoid negative values. Diffusion map embedding was subsequently applied to identify the gradient components that captured variance in the functional connectivity architecture. To enable inter-individual comparison, gradient maps were aligned across participants using iterative Procrustes rotation. The reference template for alignment was derived from the group-average functional connectivity matrix of the healthy control group [15,34].

Subsequent analyses focused on the first two dominant gradients (primary and secondary), as these components have been most reliably identified in prior work and capture clearly interpretable axes of large-scale cortical organization [35]. These two gradients were then harmonized across sites using the ComBat algorithm prior to further analyses [36].

### Statistical analysis

All group comparisons were performed on the cross-sectional imaging data acquired at study entry. To investigate alterations in connectome hierarchy from local to global scales, analyses were performed at the regional, network, and whole-brain levels for each gradient. At the regional level, parcel-wise gradient values were examined individually. At the network level, gradient values were averaged across all parcels within each canonical functional network. At the global level, whole-brain summary metrics were calculated for each gradient. Three global metrics of gradient organization were quantified for each gradient, including the variance explained ratio, the gradient range (maximum minus minimum), and gradient variation, defined as the standard deviation of gradient values [7]. Group differences among HC, CHR-C, and CHR-NC were assessed using analysis of variance (ANOVA). To account for multiple comparisons, false discovery rate (FDR) correction was applied.

### Associations between altered gradient measures and clinical variables of CHR subjects

We next examined the associations between altered gradient measures showing significant group differences and clinical symptom severity, as assessed by the four dimensions of the Scale of Prodromal Symptoms (SOPS)—positive symptoms, negative symptoms, disorganized symptoms, and general psychopathology—among all CHR participants using Pearson correlation. Additionally, we explored the correlation between these altered gradient measures and time to conversion in the CHR-C subgroup. To account for multiple comparisons, false discovery rate (FDR) correction was applied.

## Results

### Demographic and clinical characteristics

The final analytic sample for between-group comparisons included 580 participants with 90 healthy controls (HC), 56 CHR participants who converted to psychosis during follow-up (CHR-C), and 434 CHR participants who did not convert during follow-up (CHR-NC). No statistically significant differences were observed among the HC, CHR-C, and CHR-NC groups with respect to demographic characteristics, including age, gender, race, and head motion parameters. As expected, significant between-group differences were evident in baseline clinical assessment scores between CHR and HC (Table 1).

### Group comparisons of Regional gradient values

We observed two principal gradients consistent with prior reports [5,7,15]: one functional gradient spanning unimodal visual-to-sensorimotor cortices was consistently identified across all three groups, alongside one gradient conforming to the canonical sensorimotor-to-association hierarchy (Figure 1a). The visual-to-sensorimotor gradient explained on average 16.05% (SD = 1.60) of variance for all subjects, 15.99% (SD = 1.57) for HC, 16.27% (SD = 1.42) for CHR-C and 16.03% (SD = 1.62) for CHR-NC. The sensorimotor-to-association gradient explained on average 12.77% (SD = 1.25) of variance for all subjects, 12.81% (SD = 1.21) for HC, 13.16% (SD =1.12) for CHR-C and 12.71% (SD = 1.26) for CHR-NC. The relative order of these gradients, however, can vary across studies. Typically, the sensory-to-association gradient accounts for the largest proportion of variance, with the visual-to-sensorimotor gradient explaining the second largest, although some studies have reported the opposite pattern [32,37].

**Figure 1.**
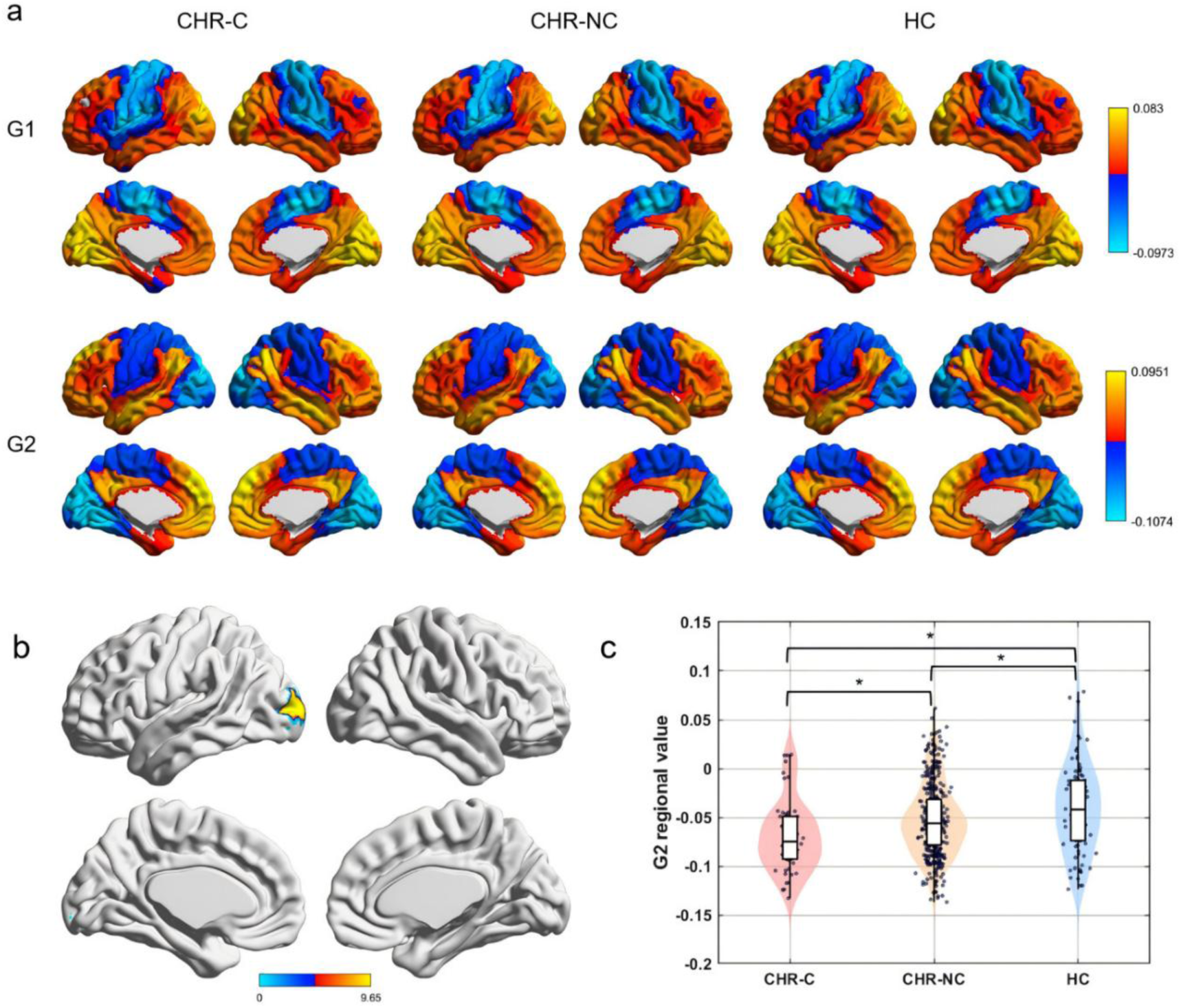
Differences of Regional-level gradient scores across groups. (a) Spatial topography of Gradient-1 and Gradient-2 across the three groups. Gradient scores are projected onto the inflated cortical surface. The top row corresponds to Gradient-1 (visual-to-sensorimotor axis) and the bottom row to Gradient-2 (sensorimotor-to-association hierarchy). Columns from left to right represent the CHR-C, CHR-NC, and HC groups, respectively. (b) Cortical surface rendering of F-values for regional sensorimotor-to-association gradient scores surviving multiple comparisons correction. (c) Violin plots with embedded boxplots and overlaid individual points showing the distribution of sensorimotor-to-association gradient scores within the significant cluster for the CHR-C, CHR-NC, and HC groups. Asterisks indicate significant post hoc differences.

Following multiple comparisons, a significant group effect was identified for sensorimotor-to-association gradient in left middle occipital gyrus (Figure 1b; F(2, 577) = 9.6501, p < 0.001, MNI coordinates: –24, –96, 6). Post hoc comparisons indicated that gradient scores in this region were significantly lower in CHR-C than in CHR-NC (p = 0.015) and HC (p < 0.001), and were also lower in CHR-NC than in HC (p = 0.008), reflecting a stepwise pattern of CHR-C < CHR-NC < HC (Figure 1c).

### Group comparisons of network gradient values

At the network level, we conducted comparative analyses of mean gradient values across the three groups using one-way ANOVA. Significant group effects were observed in three specific network domains (Figure 2a-b). The CHR-C group exhibited a significant decrease in visual-to-sensorimotor gradient scores within the Dorsal Attention Network relative to both comparison groups (F(2, 577) = 4.47, p = .012; post hoc CHR-C vs. CHR-NC: p = .028; CHR-C vs. HC: p = .010, Figure 2b1), whereas the HC and CHR-NC groups did not differ (p = .477). For sensorimotor-to-association gradient, the CHR-C group demonstrated a pronounced reduction in Visual Network scores compared to the CHR-NC group (F(2, 577) = 5.58, p = .004; post hoc CHR-C vs. CHR-NC: p = .003, Figure 2b2), with no significant differences observed between HC and CHR-NC (p = .682) and a significant difference between CHR-C and HC (p = .03). Conversely, a significant elevation in sensorimotor-to-association gradient scores was evident in the Default Mode Network for the CHR-C group relative to the CHR-NC cohort (F(2, 577) = 3.81, p = .023; post hoc CHR-C vs. CHR-NC: p = .016, Figure 2b3),with a significant increase compared to HC (p = .03), while CHR-NC and HC did not differ (p = .945).

**Figure 2.**
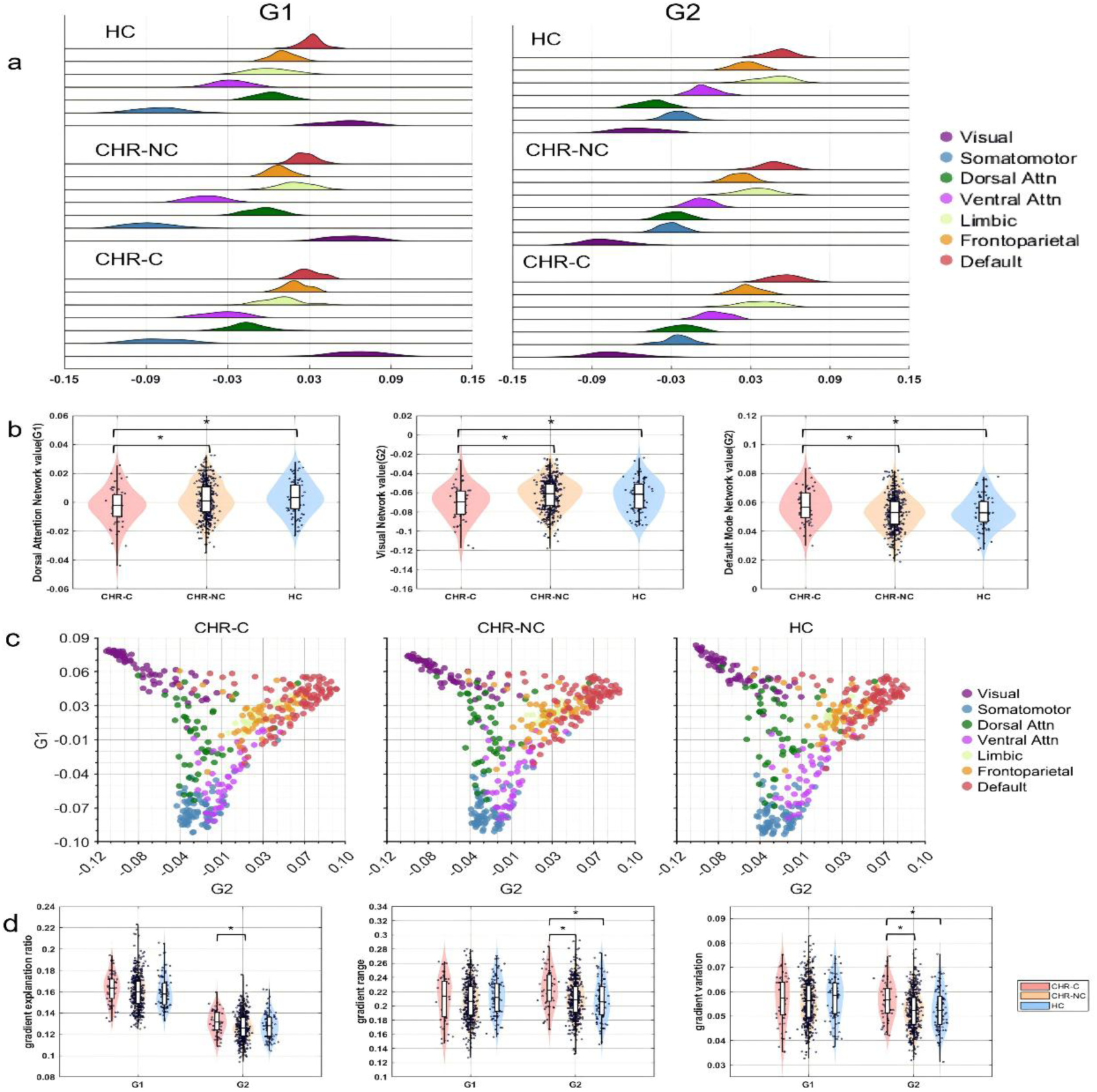
Differences of Network-level and Global-level gradient scores across groups. (a) Distribution of mean gradient scores for seven Yeo functional networks, for visual-to-sensorimotor gradient (left) and sensorimotor-to-association gradient (right)shown separately for the CHR-C, CHR-NC, and HC groups. (b) Between-group comparisons of network gradient values across the CHR-C, CHR-NC, and HC groups. b1) Differences of mean gradient scores with Dorsal Attention Network(DAN) in the visual-to-sensorimotor gradient. b2) Differences of mean gradient scores with Visual Network(VN) in the sensorimotor-to-association gradient. b3)Differences of mean gradient scores with Default Mode Network(DMN) in the sensorimotor-to-association gradient. Panels (b1-b3) display violin plots with embedded boxplots and overlaid individual points. Asterisks denote significant post hoc differences. (c) Distribution of cortical parcels in the two-dimensional gradient space defined by visual-to-sensorimotor gradient and sensorimotor-to-association gradient. Colors represent the seven Yeo functional networks. Panels from left to right correspond to the CHR-C, CHR-NC, and HC groups, respectively. (d) Between-group comparisons of global sensorimotor-to-association gradient metrics across the CHR-C, CHR-NC, and HC groups. d1)explained variance of the sensorimotor-to-association gradient. d2) range of the sensorimotor-to-association gradient. d3) variation of the sensorimotor-to-association gradient. Violin plots with embedded boxplots and overlaid individual points are shown. Asterisks denote significant post hoc differences.

### Group comparisons of Global gradient values

ANOVA revealed no significant differences in the unimodal visual-to-sensorimotor gradient among the three groups. However, analysis of the global sensorimotor-to-association gradient metrics revealed distinct alterations in the CHR-C group (Figure 2d). Specifically, the explained variance of sensorimotor-to-association gradient was significantly greater in the CHR-C group relative to the CHR-NC group (F(2, 577) = 3.20, p = .042; post hoc CHR-C vs. CHR-NC: p = .033, Figure 2d1), whereas no differences were observed between the HC group and either CHR subgroup (CHR-C vs. HC: p = .236; CHR-NC vs. HC: p = .771). Furthermore, the CHR-C group exhibited a marked expansion in sensorimotor-to-association gradient range compared to both HC and CHR-NC participants (F(2, 577) = 7.16, p < .001; post hoc CHR-C vs. CHR-NC: p < .001; CHR-C vs. HC: p = .002, Figure 2d2), with no significant difference between the HC and CHR-NC groups (p = .848). Similarly, sensorimotor-to-association gradient variation was significantly increased in the CHR-C group relative to both HC and CHR-NC cohorts (F(2, 577) = 6.08, p = .002; post hoc CHR-C vs. CHR-NC: p = .002; CHR-C vs. HC: p = .015, Figure 2d3), while HC and CHR-NC groups did not differ (p = .983). Collectively, these convergent metrics indicate a pattern of global sensorimotor-to-association gradient architecture expansion selectively expressed in individuals who subsequently converted to psychosis.

### Correlations between sensorimotor-to-association gradient metrics and clinical variables of CHR subjects

We next examined associations between sensorimotor-to-association gradient measures showing significant group differences and clinical symptom severity among CHR participants. Baseline positive symptom scores were positively correlated with both sensorimotor-to-association gradient range (r = 0.11, p = .017, Figure 3a) and sensorimotor-to-association gradient variation (r = 0.13, p = .004, Figure 3b). At the subnetwork level, positive symptom severity exhibited a negative association with sensorimotor-to-association gradient scores in the Visual Network (r = −0.11, p = .018, Figure 3c) and a positive association with sensorimotor-to-association gradient scores in the Default Mode Network (r = 0.10, p = .026, Figure 3d). Interestingly, within the CHR-C subgroup, time to conversion was negatively correlated with sensorimotor-to-association gradient explained variance (r = −0.48, p < .001, Figure 3e), indicating that individuals with lower gradient explained variance along this axis converted to psychosis more rapidly.

**Figure 3.**
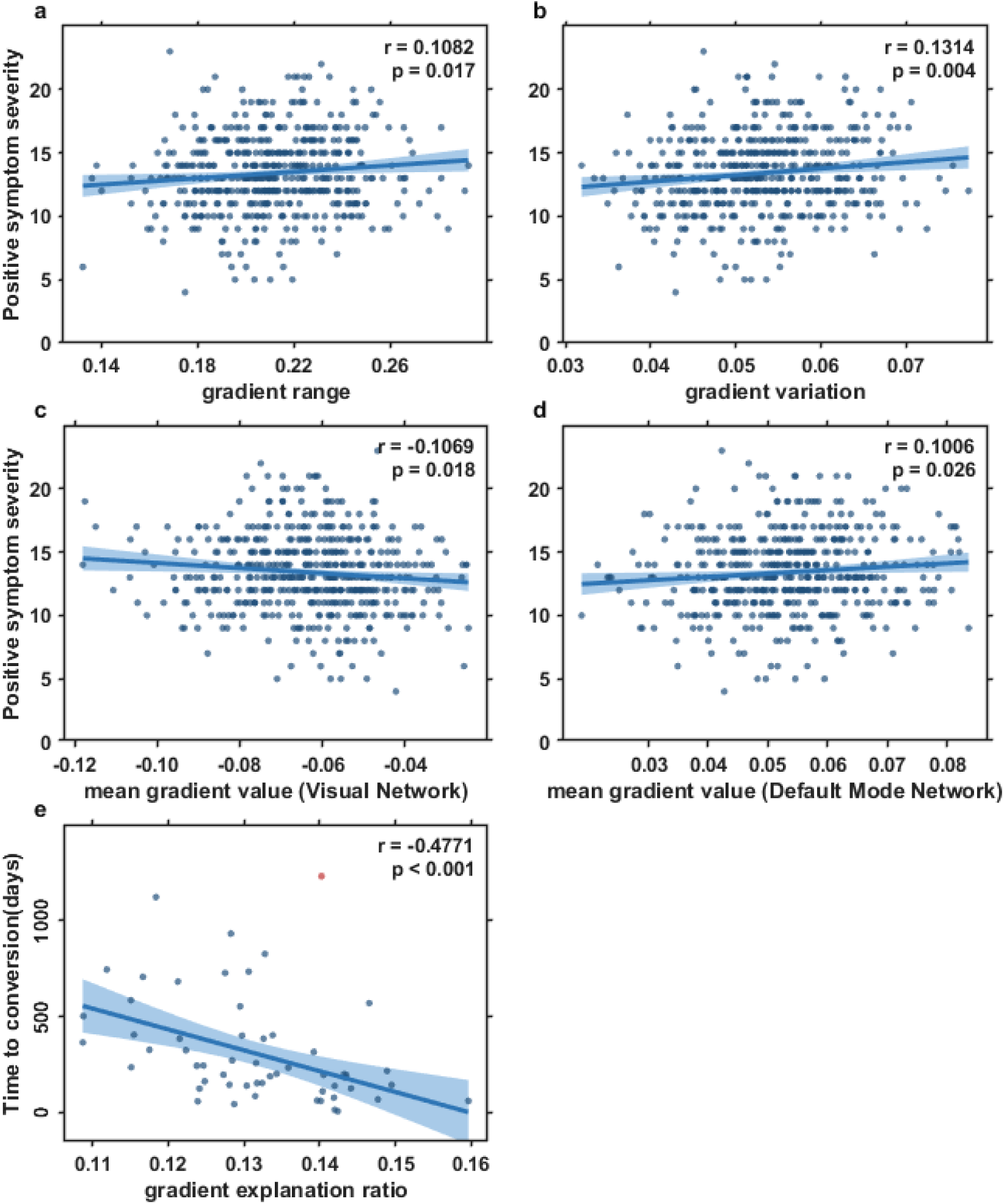
Associations between sensorimotor-to-association gradient metrics and clinical variables in CHR participants. (a) Correlation between sensorimotor-to-association gradient range and positive symptom severity. (b) Correlation between sensorimotor-to-association gradient variation and positive symptom severity. (c) Correlation between Visual Network sensorimotor-to-association gradient scores and positive symptom severity. (d) Correlation between mean gradient scores of Default Mode Network in sensorimotor-to-association gradient and positive symptom severity. (e) Correlation between sensorimotor-to-association gradient explained variance and time to conversion in the CHR-C group (outliers excluded). Each point represents an individual participant. Shaded areas denote 95% confidence intervals. Correlation coefficients (r) and p-values are displayed within each panel.

## Discussion

What pre-onset alterations in cortical hierarchy are associated with subsequent transition to psychosis in high-risk individuals? Prior work has suggested that disrupted hierarchical organization of cortical networks, particularly along the unimodal-to-transmodal axis, may represent a potential mechanistic pathway to psychotic disorders, yet direct evidence demonstrating that these large-scale hierarchical disruptions are detectable prior to psychosis onset and specifically associated with subsequent conversion remains limited. Using longitudinal outcome data from the NAPLS-3 cohort, we found that baseline alterations along the unimodal-to-transmodal gradient axis were detectable prior to psychosis onset and specifically distinguished clinical high-risk individuals who subsequently transitioned to psychosis from both non-converting high-risk participants and healthy controls, whereas non-converting individuals did not differ from healthy controls. These alterations manifested as coordinated regional, network, and global changes, reflecting a greater separation between sensory and transmodal systems and an expansion of hierarchical organization. Together, these findings identify alterations in cortical hierarchy that precede psychosis onset and are associated with subsequent transition rather than with clinical high-risk status more generally.

Notably, group differences were primarily and selectively localized to the sensorimotor-to-association gradient, suggesting that the hierarchical axis spanning unimodal sensory processing and higher-order transmodal cognition may be particularly vulnerable prior to psychosis onset. This is consistent with prior studies in schizophrenia, which have reported alterations along the sensorimotor-to-association gradient relatively consistently in both drug-naïve first-episode and chronic patients, whereas changes in other gradients, such as the visual-to-sensorimotor gradient, have been reported less consistently [4,15,16,20,21]. Our findings extend these observations by demonstrating that such hierarchical disruptions are detectable in individuals at clinical high-risk who later convert to psychosis, highlighting early sensorimotor-to-association gradient alterations as a potential neural feature linked to subsequent transition in high-risk individuals. This aligns with theoretical and computational models proposing that psychosis emerges from disrupted hierarchical information flow between bottom-up sensory inputs and top-down associative predictions, as conceptualized in hierarchical predictive processing frameworks of psychosis [38,39].

At the regional and network levels, CHR-C individuals exhibited bidirectional alterations along the sensorimotor-to-association gradient, with lower gradient values in the visual regions and the visual network and higher values in the default mode network (DMN). Because the visual system anchors the unimodal end of the cortical hierarchy and the DMN occupies its transmodal apex [13], this dissociation reflects an expanded hierarchical distance between sensory and association systems. Reduced gradient values in visual regions and the visual network align with prior findings linking visual network abnormalities to perceptual disturbances and hallucinations in schizophrenia [15,16,21], while elevated DMN values may indicate altered integration within higher-order transmodal systems supporting self-referential and abstract processing [40,41]. Overall, this pattern may reflect hierarchical decoupling, in which functional integration between bottom-up sensory input and top-down interpretative processes is weakened [42,43].

Beyond the bidirectional alterations observed in the sensorimotor-to-association gradient, we also observed changes in the visual-to-sensorimotor gradient, with reduced gradient values in the dorsal attention network (DAN) in CHR-C individuals compared to both CHR-NC and HC groups. The DAN occupies a transitional position between sensory and association systems and is critically involved in integrating sensory information with top-down attentional control [44–46]. In the context of lower visual gradient values and higher DMN gradient values, the alteration in the DAN may therefore reflect impaired coordination of attentional resources across hierarchical levels, consistent with the notion of hierarchical decoupling. Such dysfunction in intermediate networks may contribute to aberrant salience processing, a core feature of psychosis [47], indicating that hierarchical disruptions extend beyond the extreme ends of the cortical gradient and involve transitional systems that support cross-level integration.

At the global level, CHR-C individuals exhibited increased explained variance, range, and variation of the sensorimotor-to-association gradient, indicating an expansion of hierarchical organization. While prior studies in established schizophrenia have reported gradient compression [15,20], our findings suggest that individuals who subsequently convert to psychosis may be characterized by a distinct pattern of global reorganization prior to onset [48,49]. Notably, the explained variance of the sensorimotor-to-association gradient was negatively associated with time to conversion, such that individuals with higher explained variance transitioned more rapidly to psychosis. This suggests that increased hierarchical dominance along the unimodal-to-transmodal axis may reflect early maladaptive reorganization, potentially constraining the brain’s functional architecture and exaggerating the functional separation between sensory and transmodal systems. Such organization is consistent with reduced integration across processing levels and greater imbalance between bottom-up sensory information and top-down predictive or interpretive processes, as proposed in hierarchical models of psychosis [38,39,50].

The clinical relevance of these findings is further underscored by their associations with symptom severity. Across CHR individuals, greater global sensorimotor-to-association gradient range and variation were associated with higher positive symptom severity, indicating that hierarchical expansion is linked to subthreshold psychotic experiences. At the network level, the opposing associations observed in the visual network (negative) and DMN (positive) mirrored the group-level differences in CHR-C participants, further supporting an association between greater hierarchical separation of sensory and transmodal systems and symptom severity. Such a pattern aligns with models of aberrant perceptual inference [38], in which disrupted integration between bottom-up sensory input and top-down predictions leads to misinterpretation of environmental stimuli and emergence of psychotic symptoms.

Several limitations should be noted. First, although the study included a large multisite sample of individuals at clinical high risk for psychosis, the number of participants who subsequently transitioned to psychosis remained moderate (N = 56), and the marked imbalance in group sizes may have affected statistical power and precision. Replication in larger independent high-risk cohorts will therefore be important to establish the robustness and generalizability of these findings. Second, neuroimaging data were acquired only at baseline. Although clinical outcomes were assessed longitudinally, the present design cannot directly characterize within-individual changes in cortical hierarchy across the transition to psychosis, and variation in age and time to conversion may influence the observed associations.

Third, medication exposure remains a potential confound. Although chlorpromazine-equivalent dose did not differ significantly between CHR-C and CHR-NC groups and was not associated with the gradient measures examined (all p > 0.50), residual medication effects cannot be fully excluded. Fourth, the analyses focused on the cerebral cortex, whereas subcortical and cerebellar regions, including the thalamus, hippocampus, and cerebellum, also play important roles in psychosis pathophysiology [51,52]. Future work incorporating these systems may provide a more comprehensive characterization of hierarchical reorganization associated with psychosis risk.

### Ethical Standards

The NAPLS-3 study protocols were approved by the institutional review boards of all participating institutions. The present study used de-identified publicly available data from the NAPLS-3 database. Analyses conducted in the present work were also approved by the Ethics Committee of Southwest University (approval number H25055). The authors assert that all procedures contributing to this work comply with the ethical standards of the relevant national and institutional committees on human experimentation and with the Helsinki Declaration of 1975, as revised in 2013.

### Funding

This work was supported by the National Natural Science Foundation of China (Grant No’s. 32300861 and 82202247) and Natural Science Foundation of Chongqing (CSTB2023NSCQ-MSX0896).

### Author Contribution

Yulin Wang and Debo Dong conceived and designed the study. Yichuan Wang, Shaolei Guo, and Anfu Deng performed the analyses. Bihua Xu and Jiaxi Liao contributed to visualization of the results. Yichuan Wang, Enhui Zhang, Yulin Wang, and Jiaxi Liao interpreted the results. Yichuan Wang, Enhui Zhang, and Debo Dong wrote the original manuscript; Yulin Wang and Debo Dong supervised the study and obtained funding. All authors contributed to manuscript revision and approved the final version of the manuscript.

### Data Availability

The NAPLS-3 data are available to qualified researchers through the NIMH Data Archive (NDA). The authors do not have permission to distribute these data directly. Details of the study design and data collection procedures have been described previously [27].

### Analytic Code Availability

Connectome gradient mapping was implemented using the open-source BrainSpace toolbox (version 0.1.1), which is freely available at https://github.com/MICA-MNI/BrainSpace under a BSD 3-Clause license. Custom MATLAB and Python scripts used for data processing and statistical analyses are not publicly available.

